# Rates of Adherence to Colorectal Cancer Medications and Predictors of Non-Adherence: A Systematic Review and Meta-Analysis

**DOI:** 10.64898/2026.01.07.26343638

**Authors:** Ruchika Raj, Tadesse M. Abegaz, Regina N. Nechi, Macarius Donneyong

**Author notes:** Corresponding author information: Name: Macarius Donneyong, PhD, MPH, Affiliation: The Ohio State University Division of Pharmacy Practice and Science Email address, Twitter handle: N/A. List of where and when the study has been presented elsewhere: 2024 New York University Martin Luther King Jr. Undergraduate Research Conference 2025 Ohio State University College of Pharmacy Summer Undergraduate Research Fellow Conference. The authors declare no competing interests.

## Abstract

**PURPOSE:** To synthesize adherence rates to colorectal cancer medications and identify predictors of nonadherence.

**METHODS:** Following PRISMA, we searched PubMed, Embase, PsycINFO, and Web of Science through August 13, 2024. Observational studies reporting adherence or predictors were eligible. Two reviewers independently screened and extracted data and assessed risk of bias using JBI’s Checklist for Cohort Studies. Adherence was grouped by measurement approach: claims-based PDC/MPR, chart/clinical record review, or patient-reported. Random-effects meta-analyses were performed within clinically and methodologically homogeneous subgroups.

**RESULTS:** Thirteen studies (n = 13) met inclusion, with adherence ranging from 33% to 100%. In claims-based analyses using PDC/MPR thresholds, pooled adherence was about 40% [95% CI, 0.36-0.44] with substantial heterogeneity (I^2^ = 84.6%). Pooled adherence was about 83% in both chart/record [95% CI, 0.44-0.97] (I^2^ = 71.4%) or patient-reported measures [95% CI, 0.68-0.92] (I^2^ = 93.8%), also with substantial heterogeneity. Nonadherence was more likely with advanced stage, ECOG ≥1, multiple prior regimens, female sex, and treatment-related adverse events. The overall risk of bias was low, although some included studies lacked complete follow-up or strategies to address it.

**CONCLUSION:** We synthesized adherence to CRC medications and identified consistent predictors of nonadherence. Adherence was lowest with claims-based PDC/MPR and higher with chart or patient-reported measures. These findings support targeted interventions for patients at higher risk of non-adherence, including those in the advanced stage of the disease, those with multiple regimens, and those experiencing adverse events. Future work should use standardized adherence definitions and metric-specific reporting to enable valid pooling.

## INTRODUCTION

Colorectal cancer is among the most common cancers worldwide, with 1,926,425 new cases reported in 2022.^1^ In the United States, excluding skin cancers, it is the third most frequently diagnosed cancer in both men and women.^2^ Among the ten most prevalent cancers globally, colorectal cancer has the highest proportion of patients receiving chemotherapy (70.3%).^3^ Because chemotherapy is central to colorectal cancer treatment, its effectiveness relies heavily on patients’ adherence to prescribed regimens. Poor adherence can compromise treatment efficacy, resulting in poor prognosis and increased medical costs. ^4^

Despite the importance of maintaining adherence, evidence on adherence rates and predictors of nonadherence in colorectal cancer remains scattered. To our knowledge, no prior systematic reviews or meta-analyses have synthesized this information. The objectives of this study are to estimate current adherence rates to colorectal cancer medications, identify predictors of nonadherence, and provide pooled estimates to inform clinical practice and the development of future interventions.

## METHODS

This review was conducted and reported following the PRISMA (Preferred Reporting Items for Systematic Reviews and Meta-Analyses) guidelines.

### Eligibility Criteria

Studies were eligible for inclusion if they examined adherence to chemotherapy among adults diagnosed with colorectal, colon, or rectal cancer. We included cohort studies that reported adherence rates or evaluated predictors of adherence or nonadherence. We excluded studies focused exclusively on non-chemotherapy medications, reviews or meta-analyses, and studies that did not report adherence outcomes. Articles not published in English or those lacking original, patient-level adherence data were also excluded.

### Information Sources

We searched PubMed, PsycINFO (via Ovid), Embase (via Ovid), and Web of Science to identify studies. The search was conducted from May 1, 2024, to August 13, 2024.

### Study Selection

Each of the two reviewers (RR and TA) independently reviewed studies identified using relevant search terms from the four electronic databases for relevance using predetermined inclusion and exclusion criteria, first by title and abstract then by full text. A detailed list of search terms used to identify studies is included in the attached supplementary file (Section A). Discrepancies in relevance decisions were resolved by a meeting of the two reviewers, where a final decision was reached. Covidence Review Management Software was used to organize the process. Two reviewers (RR and TA) independently screened all identified records using prespecified inclusion and exclusion criteria. Screening was conducted in two stages: title/abstract review followed by full-text assessment. The full search strategy is provided in Supplementary Section A. Discrepancies were resolved through discussion until consensus was reached. Covidence software was used to facilitate study screening and workflow management.

### Data Extraction

Two reviewers (RR and TA) independently extracted information from each including study using a standardized form. Extracted study characteristics included study ID, substudy, adherence definition, adherence measurement method, chemotherapy regimen, type of adherence variable, adherence rate (for continuous measures), proportion or number adherent (for dichotomous measures), sample size, country, study design, and year of publication.

Data on predictors of nonadherence were also collected. These included the predictor category, variable type (categorical, binary, or continuous), the effect measure reported in the study, the direction of association, confidence intervals, p-values, and whether estimates were adjusted. Individual effect sizes (e.g., ORs or HRs) were extracted for completeness but were not tabulated, as they are not directly comparable across heterogeneous study designs.

### Synthesis of Results

Studies were grouped into three categories based on the **source of data** used to assess adherence: **Administrative/Claims Data**, **Clinical Record Review**, and **Patient-Reported/Direct Measures**. The Administrative/Claims Data group included studies that relied on dispensing records, insurance claims, or other large administrative databases. The Clinical Record Review group comprised studies assessing adherence through medical chart review, clinician documentation, or pharmacist verification of completed treatment cycles or doses. The Patient-Reported/Direct Measures group included studies that used self-report instruments, patient questionnaires, or direct methods such as pill counts.

Continuous adherence metrics (e.g., Proportion of Days Covered [PDC], Medication Possession Ratio [MPR]) were dichotomized when possible using data reported in the original studies. Studies presenting adherence exclusively as a continuous variable that could not be reliably dichotomized were excluded from quantitative synthesis. For each subgroup, data were tabulated and analyzed separately using the meta package in RStudio. Random-effects models were applied to compute pooled estimates, and I^2^ statistics were calculated to assess statistical heterogeneity. Study characteristics were then reviewed to explore potential sources of heterogeneity. Sensitivity analyses were not performed.

### Risk of Bias Assessment

The risk of bias for each included study was evaluated using the Joanna Briggs Institute (JBI) Critical Appraisal Checklist for Cohort Studies. The checklist assesses potential sources of bias across domains including participant selection, measurement of exposure and outcomes, identification and management of confounding factors, and adequacy of follow-up.Each criterion was rated as “yes,” “no,” “unclear,” or “not applicable,” and overall study quality was determined based on the number of criteria met by assigning numerical values to each of the checklist item responses (“Yes” = 1, “No,” “Unclear” = 0, “Not applicable” = excluded from checklist) and computing a total score by adding the total number of “Yes” responses and dividing by total number of applicable checklist items. Quality was then categorized as High(≥80%), moderate (60–79%), or low (< 60%) for synthesis.

## RESULTS

The literature search and selection process are described in Figure 1. The database search yielded 2,152 records, of which 1,023 duplicates were removed. Among the remaining 1,129 unique studies, 1,040 were excluded during title and abstract screening. The full texts of 72 articles were reviewed in detail, and 59 were excluded for the following reasons: case study (n = 1), wrong disease (n = 4), review article (n = 1), inclusion of other cancer types (n = 8), full text unavailable (n = 33), full text not available in English (n = 3), not focused on medication adherence (n = 3), neoadjuvant therapy (n = 2), or failure to report adherence rates or predictors (n = 4).

**Figure 1:**
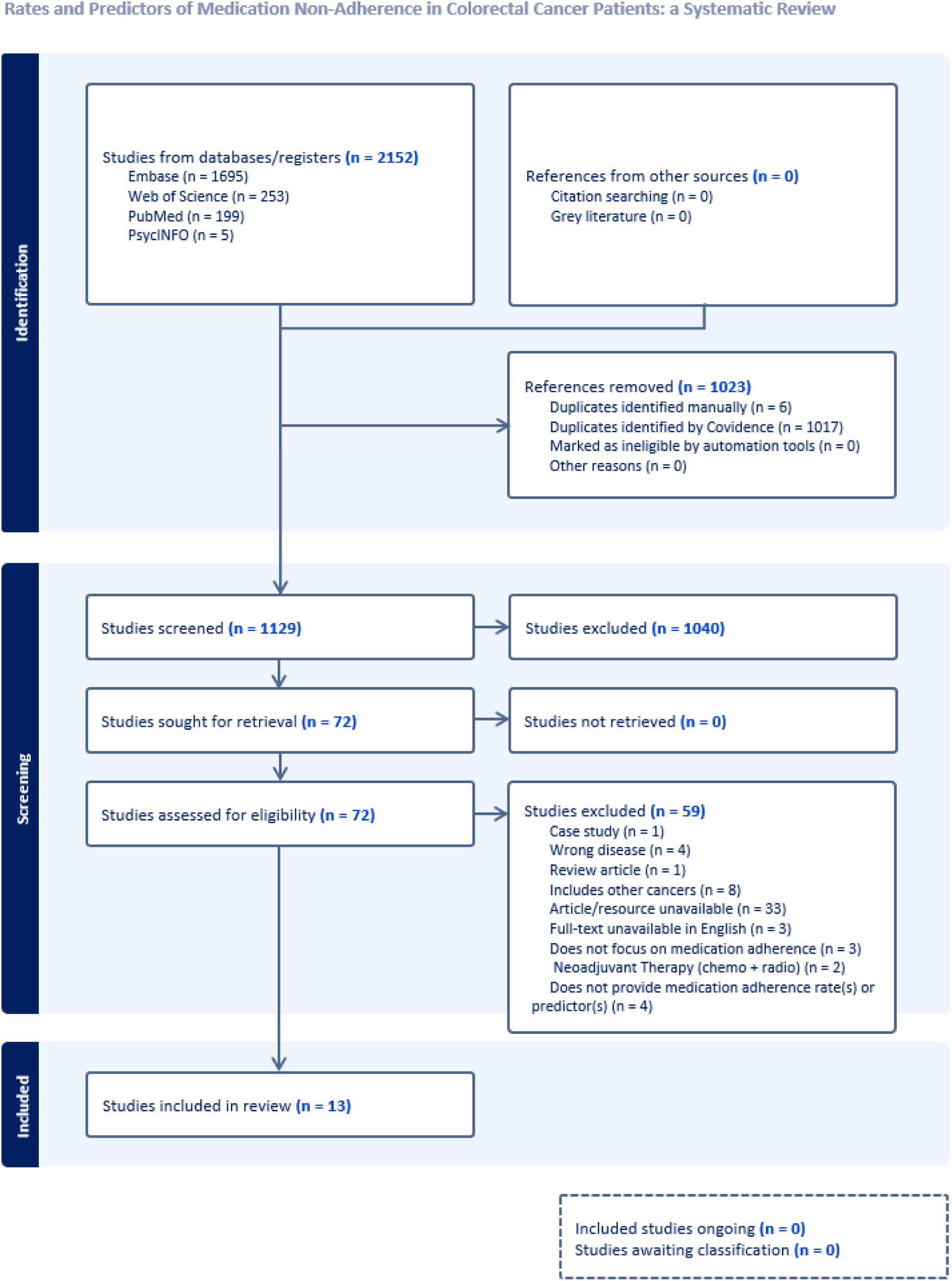
PRISMA flow diagram.

**Figure 1:**
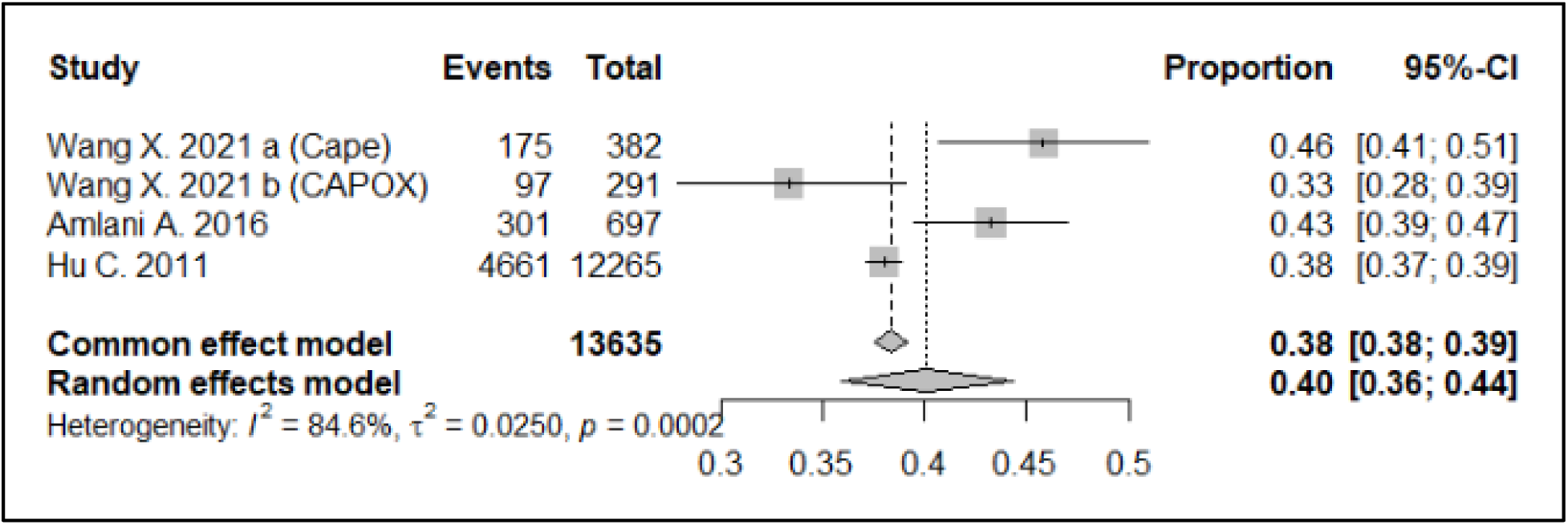
Forest Plot of “Administrative/Claims Data” Studies. This forest plot summarizes adherence rates for chemotherapy regimens in patients with colorectal cancer, as reported in studies primarily utilizing administrative or claims data to calculate adherence.

**Figure 2:**
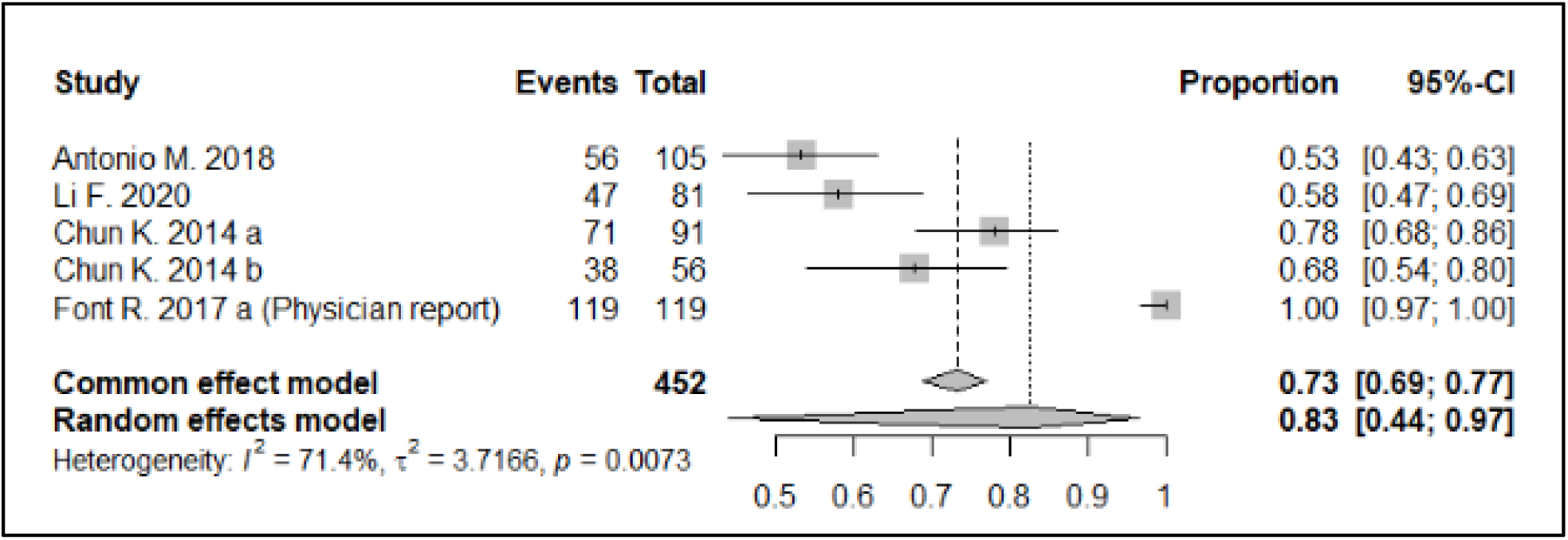
Forest Plot of “Clinical Record Review” Studies. This forest plot summarizes adherence rates for chemotherapy regimens in patients with colorectal cancer, as reported in studies primarily utilizing data from patient medical charts, physician notes, or pharmacist verification of treatment cycles/doses completed to calculate adherence.

**Figure 3:**
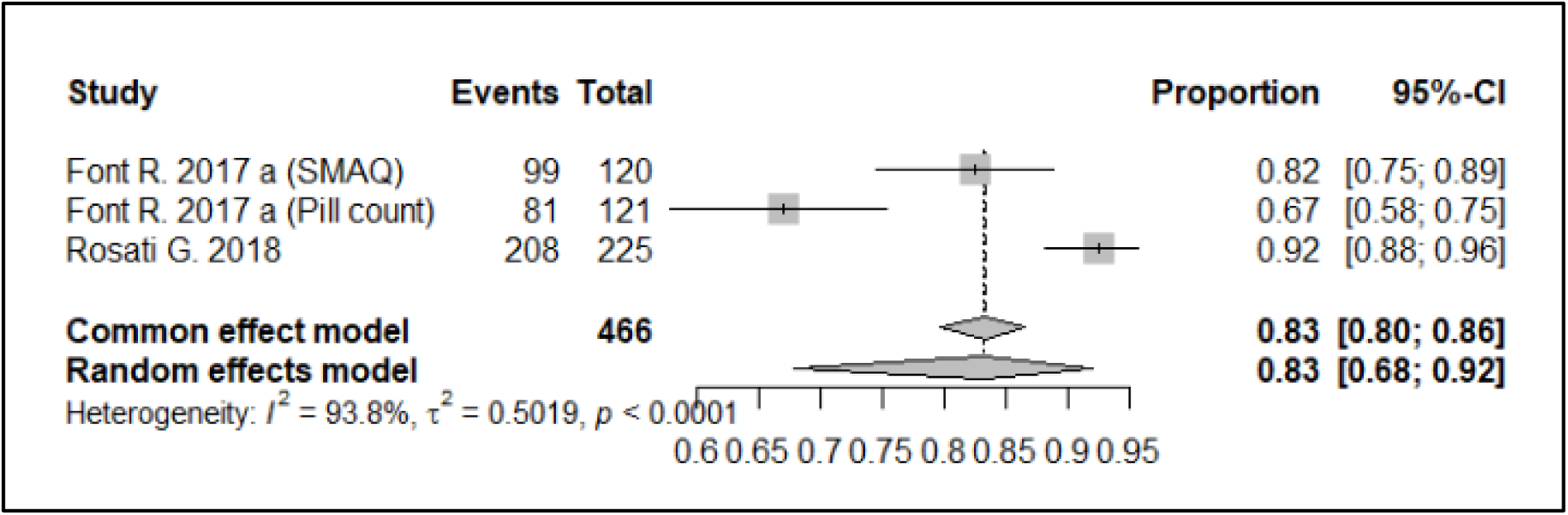
Forest Plot of “Patient-Reported/Direct Measures” Studies. This forest plot summarizes adherence rates for chemotherapy regimens in patients with colorectal cancer, as reported in studies primarily utilizing data from patient questionnaires, direct pill counts, or patient self-report to calculate adherence.

**Figure 4:**
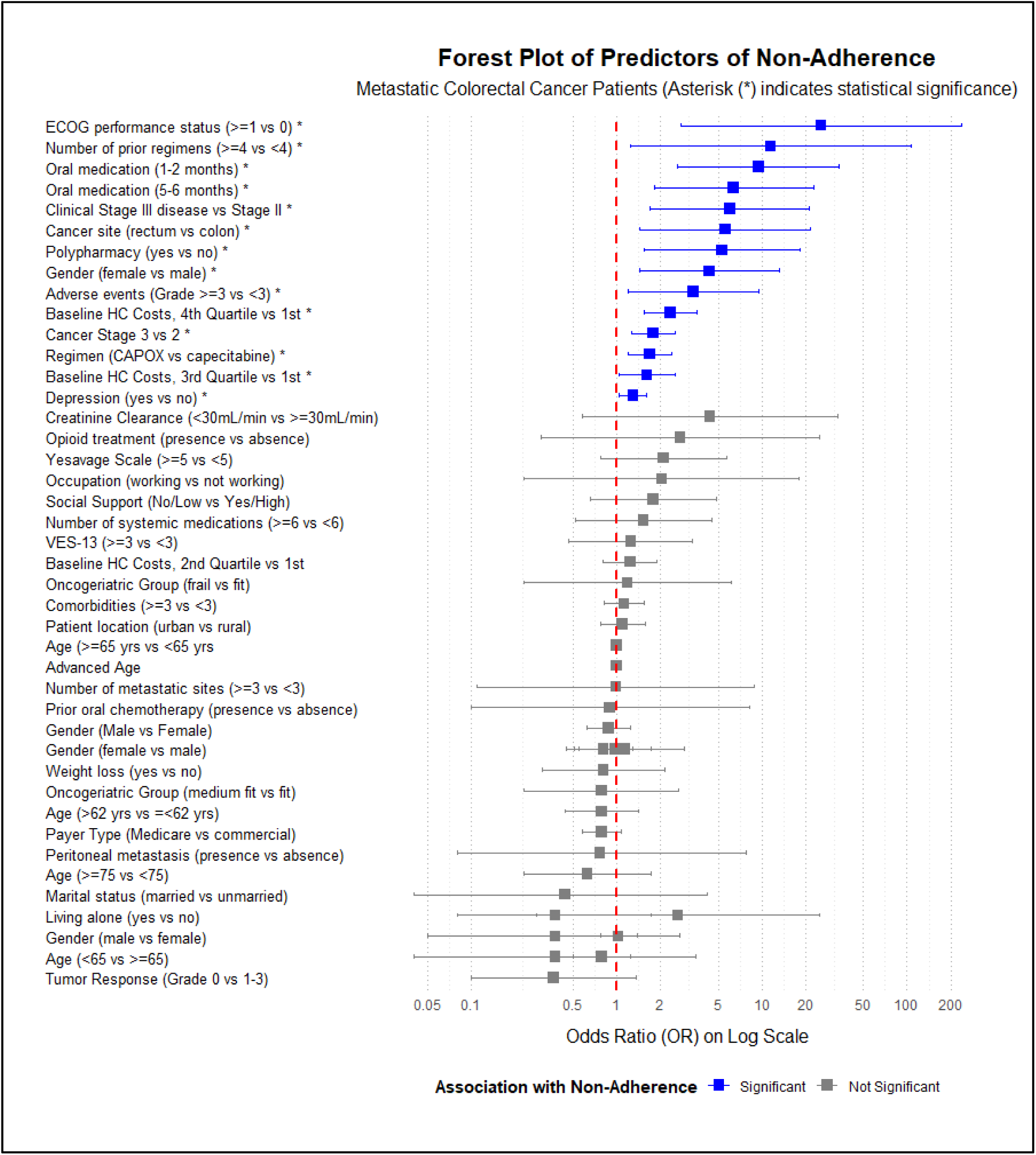
Forest Plot of Predictors of Non-Adherence in Patients with Colorectal Cancer. This forest plot visualizes odds ratios and confidence intervals for both significant and non-significant predictors of non-adherence. A predictor was considered statistically significant if its p-value was less than 0.05 and the lower bound of its confidence interval was greater than an odds ratio of 1. Certain predictors have two plotted odds ratios as the same predictor was cited in multiple included studies.

A total of 13 studies met the eligibility criteria and were included in this review. The included studies were published between 2011 and 2021 and conducted across six countries: Japan, Italy, Spain, the United States, Canada, and the Republic of Korea. Most studies employed retrospective or prospective observational designs, with sample sizes ranging from 34 to 12,265 participants. The chemotherapy agents assessed included regorafenib, capecitabine, oxaliplatin-based regimens (eg, CAPOX, FOLFOX, FOLFOX6, XELOX), irinotecan-based regimens, trifluridine/tipiracil, and 5-fluorouracil (5-FU) combinations.

Study quality, as assessed using the Joanna Briggs Institute (JBI) Critical Appraisal Checklist for Cohort Studies, ranged from 33.3% to 100%. Three studies were rated as high quality (Font et al., 2017; Kawakami et al., 2019; Amlani et al., 2016), eight as moderate quality (DelPrete et al., 2017; Antonio et al., 2018; Nagamatsu et al., 2019; Li et al., 2020; Sugita et al., 2016; Rosati et al., 2018; Wang et al., 2021; Hu et al., 2011), and two as low quality (Kawakami et al., 2015; Chun et al., 2014).

Key characteristics of the included studies are summarized in Table 1.

**Table 1.**
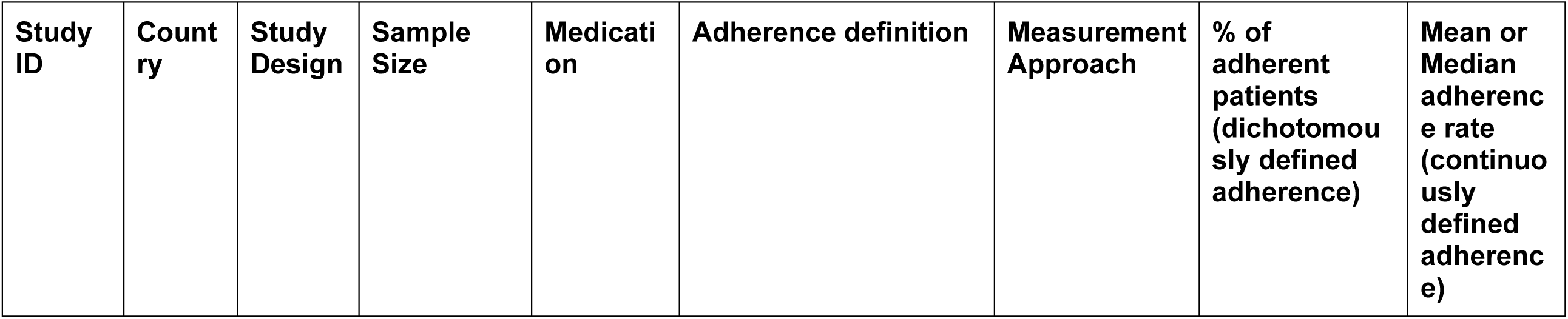
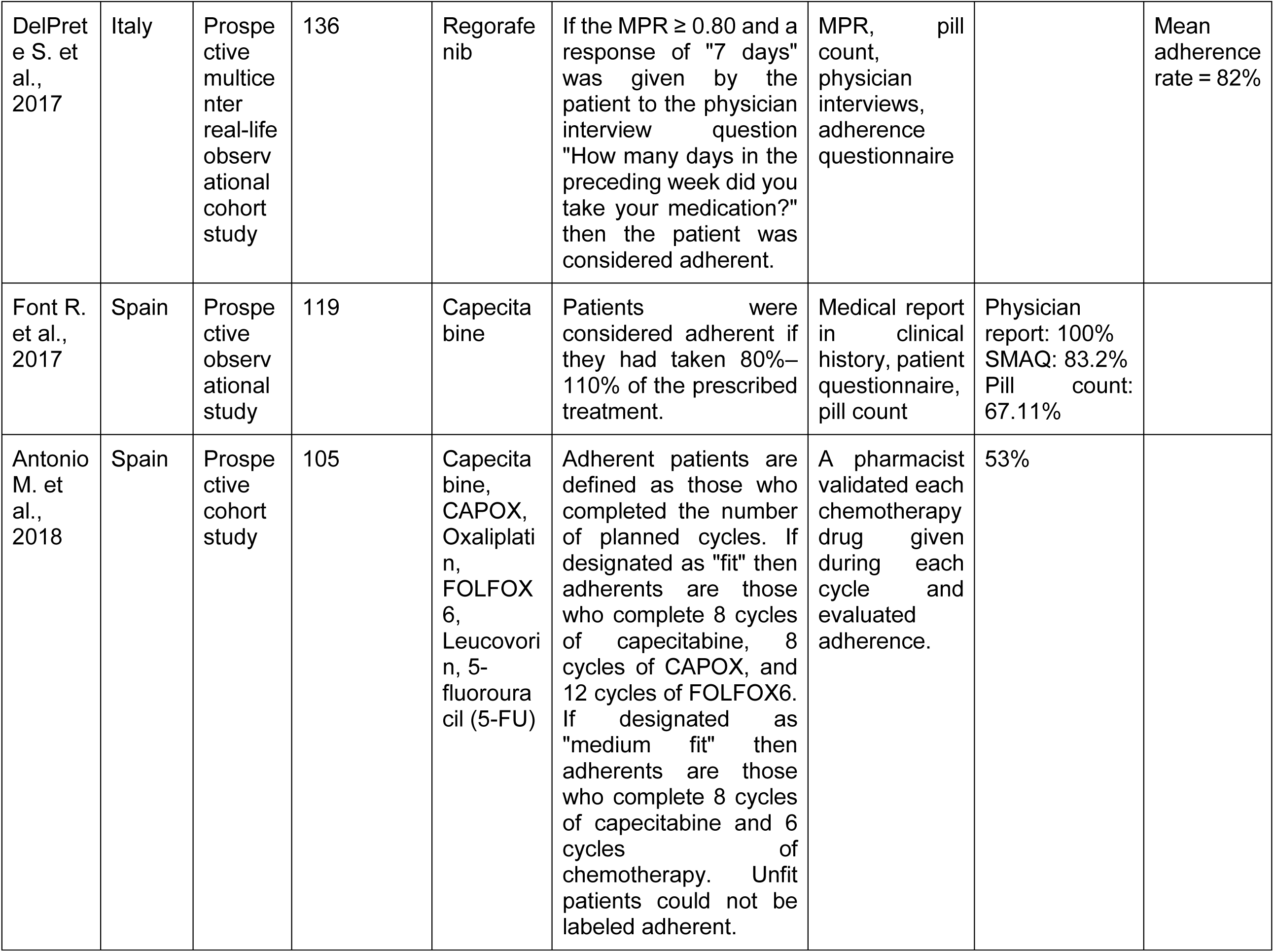
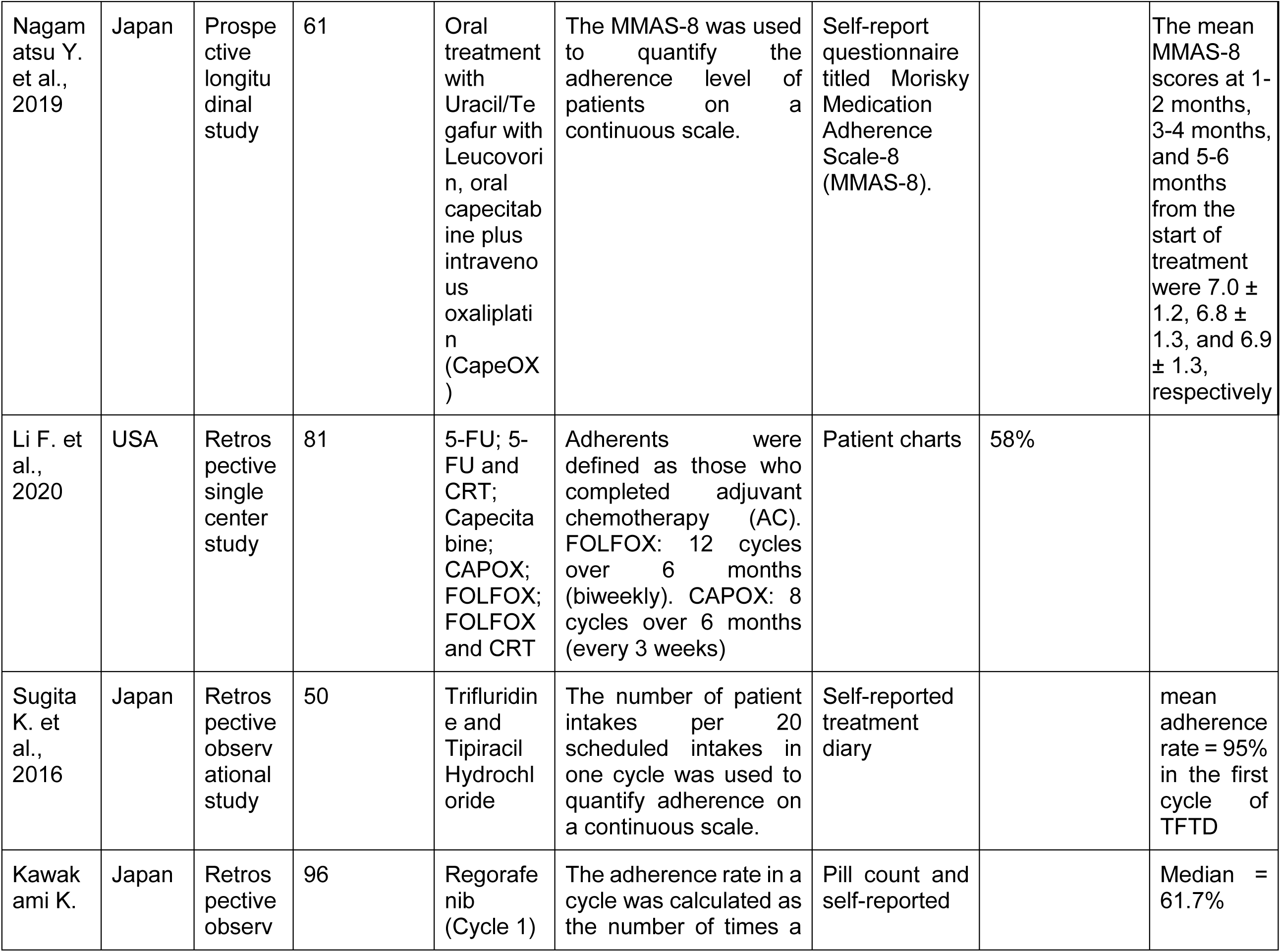
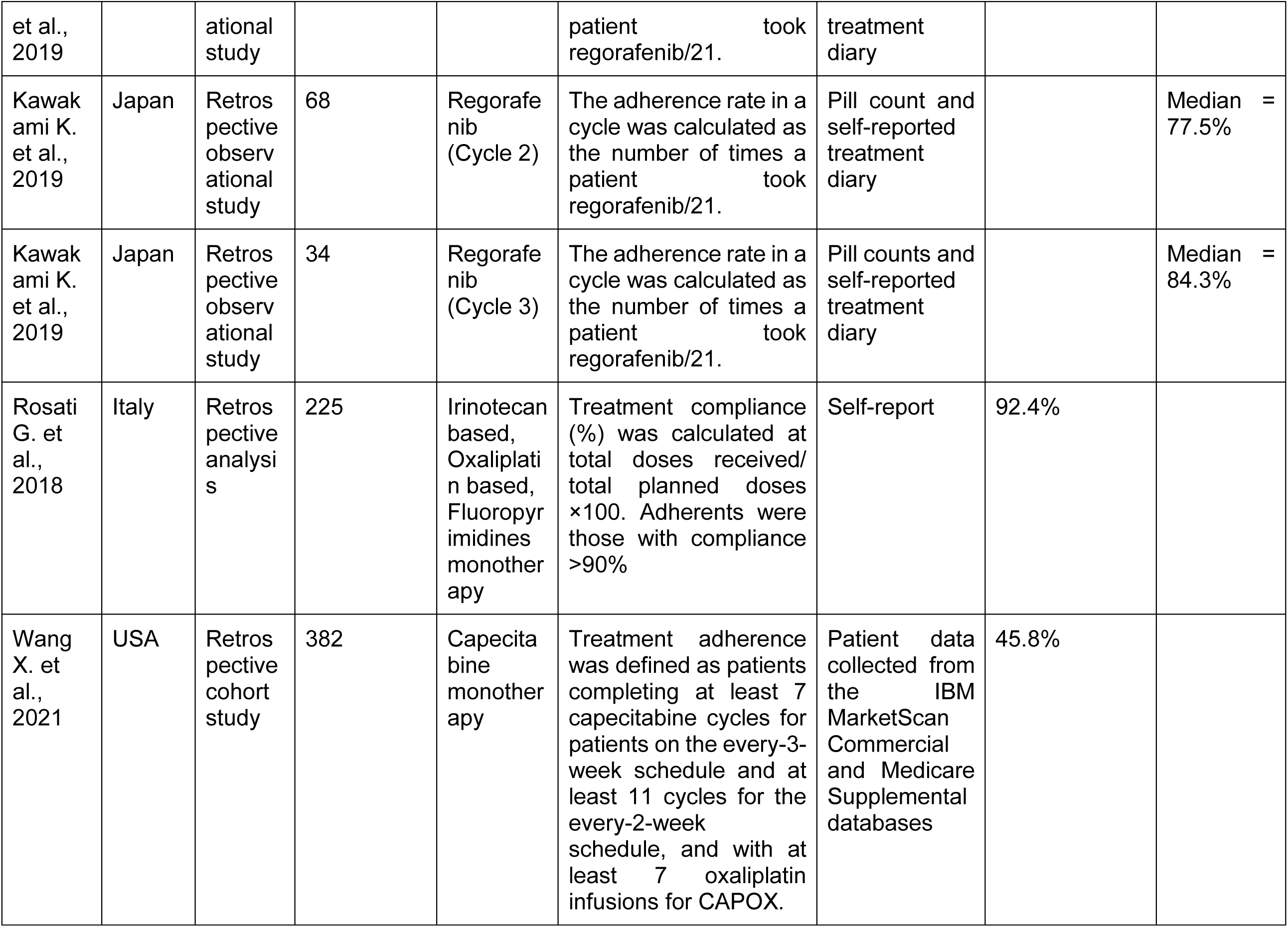
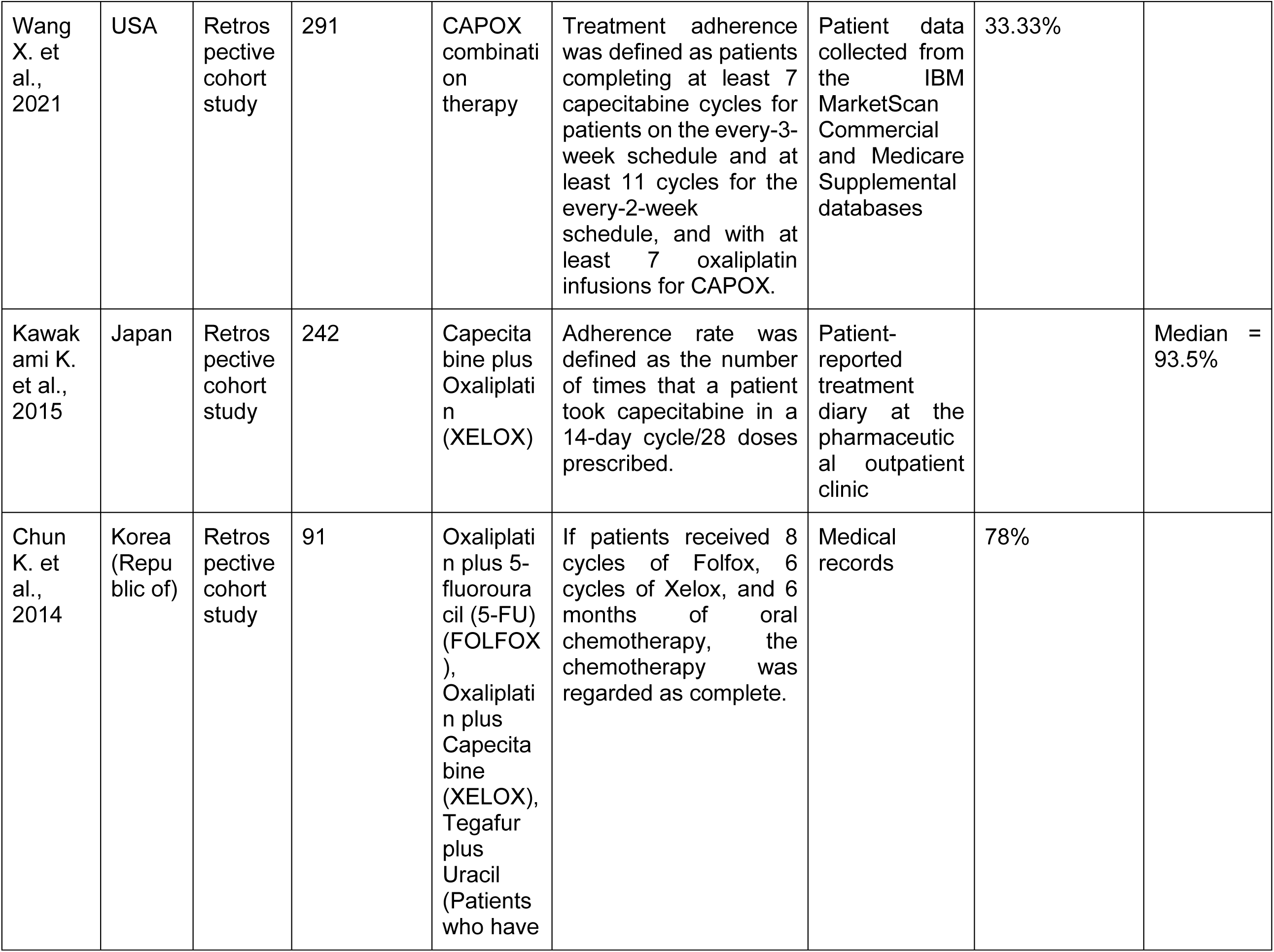
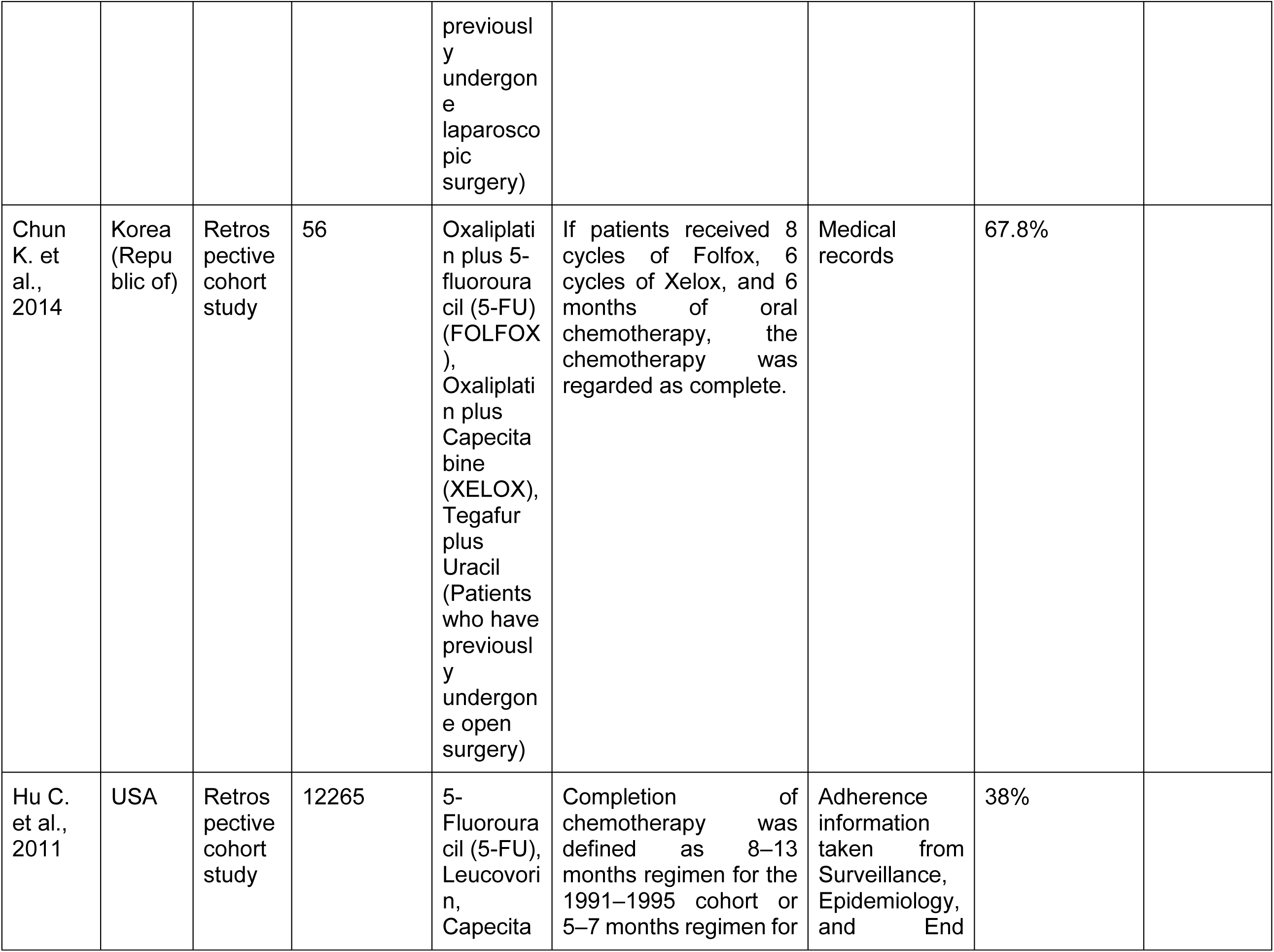
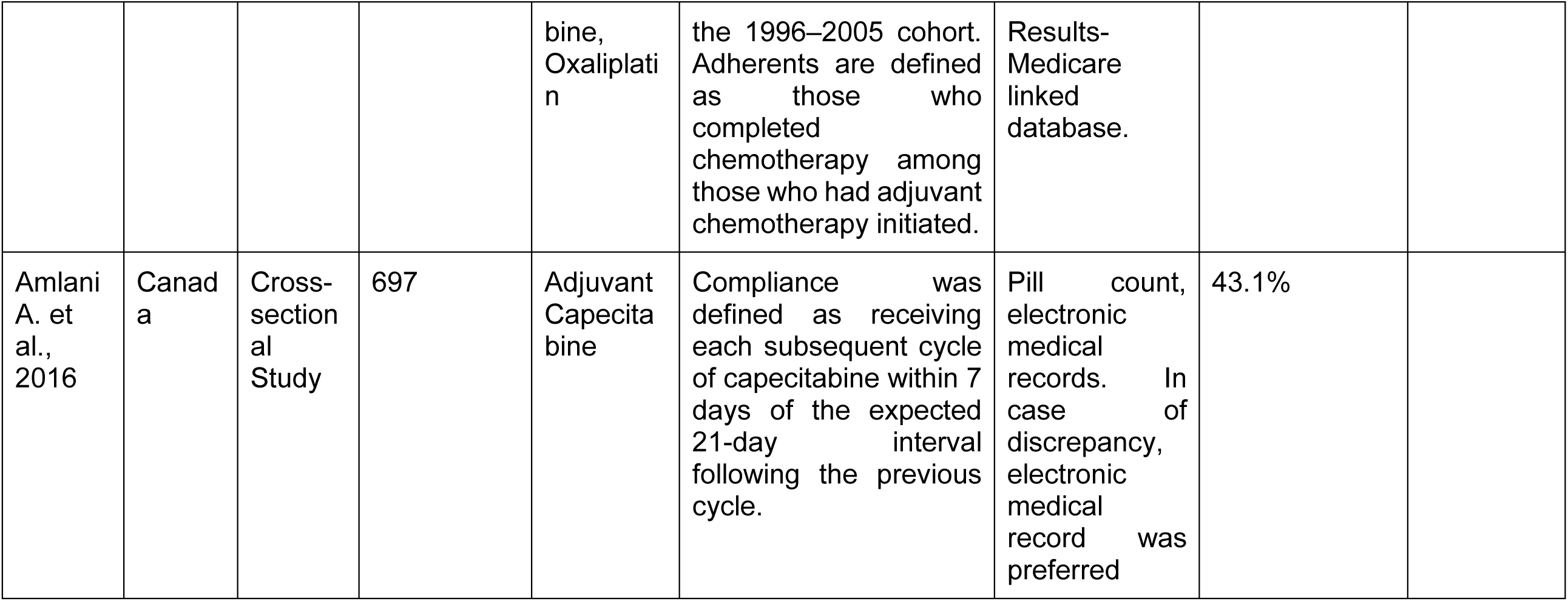
Study General Characteristics.

### Predictors of Nonadherence

Across the included studies, several patients, clinical, and treatment-related factors emerged as significant predictors of nonadherence to colorectal cancer chemotherapy. Predictors most frequently associated with a higher likelihood of nonadherence included advanced cancer stage, poorer performance status (ECOG ≥1), polypharmacy, rectal tumor site, female sex, multiple prior chemotherapy regimens, severe adverse events, and certain psychological or functional characteristics. In contrast, factors such as living alone, higher HADS scores, and lower nausea and vomiting scores tended to be associated with better adherence in some cohorts. Tables 2A and 2B summarize the key factors linked to higher and lower odds of nonadherence across studies.

**Table 2A.**
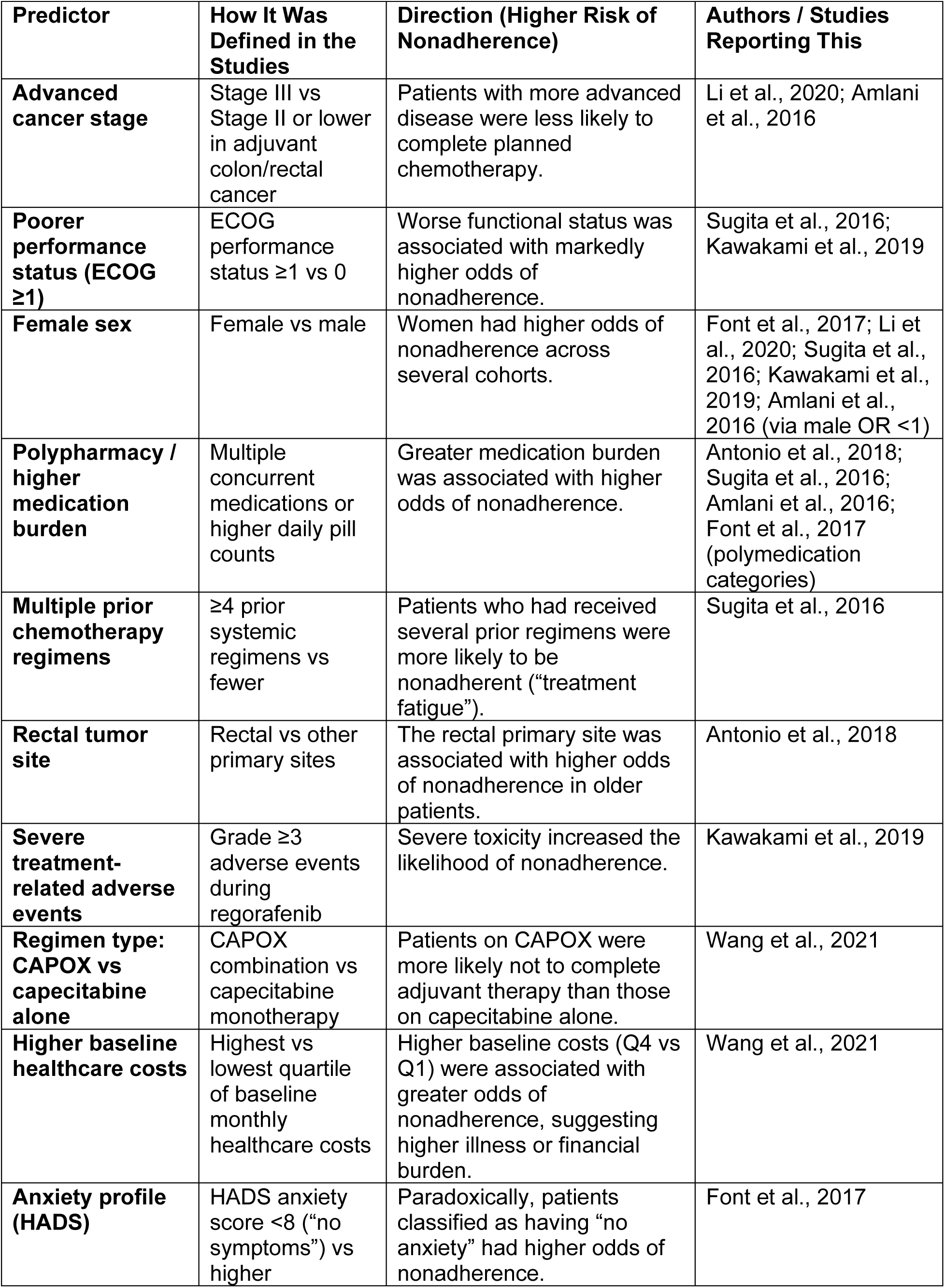
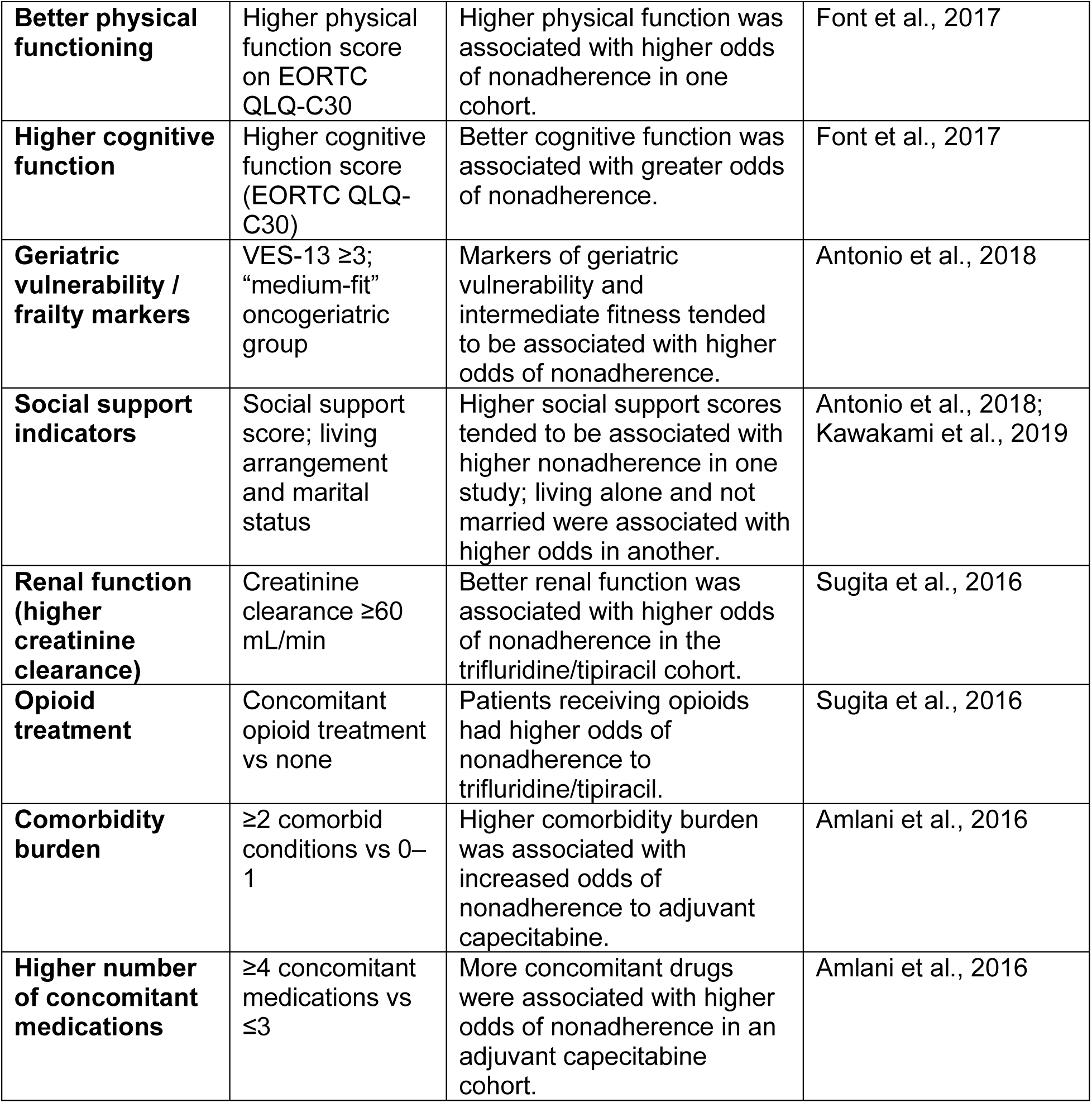
Predictors Associated With Higher Risk of Nonadherence to Colorectal Cancer Chemotherapy.

ADHERENCE TO COLORECTAL CANCER MEDICATION
| Predictor | How It Was Defined in the Studies | Direction (Higher Risk of Nonadherence) | Authors / Studies Reporting This |
| --- | --- | --- | --- |
| **Advanced cancer stage** | Stage III vs Stage II or lower in adjuvant colon/rectal cancer | Patients with more advanced disease were less likely to complete planned chemotherapy. | Li et al., 2020; Amlani et al., 2016 |
| **Poorer performance status (ECOG $\geq 1$)** | ECOG performance status $\geq 1$ vs 0 | Worse functional status was associated with markedly higher odds of nonadherence. | Sugita et al., 2016; Kawakami et al., 2019 |
| **Female sex** | Female vs male | Women had higher odds of nonadherence across several cohorts. | Font et al., 2017; Li et al., 2020; Sugita et al., 2016; Kawakami et al., 2019; Amlani et al., 2016 (via male OR $<1$) |
| **Polypharmacy / higher medication burden** | Multiple concurrent medications or higher daily pill counts | Greater medication burden was associated with higher odds of nonadherence. | Antonio et al., 2018; Sugita et al., 2016; Amlani et al., 2016; Font et al., 2017 (polymedication categories) |
| **Multiple prior chemotherapy regimens** | $\geq 4$ prior systemic regimens vs fewer | Patients who had received several prior regimens were more likely to be nonadherent ("treatment fatigue"). | Sugita et al., 2016 |
| **Rectal tumor site** | Rectal vs other primary sites | The rectal primary site was associated with higher odds of nonadherence in older patients. | Antonio et al., 2018 |
| **Severe treatment-related adverse events** | Grade $\geq 3$ adverse events during regorafenib | Severe toxicity increased the likelihood of nonadherence. | Kawakami et al., 2019 |
| **Regimen type: CAPOX vs capecitabine alone** | CAPOX combination vs capecitabine monotherapy | Patients on CAPOX were more likely not to complete adjuvant therapy than those on capecitabine alone. | Wang et al., 2021 |
| **Higher baseline healthcare costs** | Highest vs lowest quartile of baseline monthly healthcare costs | Higher baseline costs (Q4 vs Q1) were associated with greater odds of nonadherence, suggesting higher illness or financial burden. | Wang et al., 2021 |
| **Anxiety profile (HADS)** | HADS anxiety score $<8$ ("no symptoms") vs higher | Paradoxically, patients classified as having "no anxiety" had higher odds of nonadherence. | Font et al., 2017 |
| **Better physical functioning** | Higher physical function score on EORTC QLQ-C30 | Higher physical function was associated with higher odds of nonadherence in one cohort. | Font et al., 2017 |
| **Higher cognitive function** | Higher cognitive function score (EORTC QLQ-C30) | Better cognitive function was associated with greater odds of nonadherence. | Font et al., 2017 |
| **Geriatric vulnerability / frailty markers** | VES-13 $\geq 3$; "medium-fit" oncogeriatric group | Markers of geriatric vulnerability and intermediate fitness tended to be associated with higher odds of nonadherence. | Antonio et al., 2018 |
| **Social support indicators** | Social support score; living arrangement and marital status | Higher social support scores tended to be associated with higher nonadherence in one study; living alone and not married were associated with higher odds in another. | Antonio et al., 2018; Kawakami et al., 2019 |
| **Renal function (higher creatinine clearance)** | Creatinine clearance $\geq 60$ mL/min | Better renal function was associated with higher odds of nonadherence in the trifluridine/tipiracil cohort. | Sugita et al., 2016 |
| **Opioid treatment** | Concomitant opioid treatment vs none | Patients receiving opioids had higher odds of nonadherence to trifluridine/tipiracil. | Sugita et al., 2016 |
| **Comorbidity burden** | $\geq 2$ comorbid conditions vs 0–1 | Higher comorbidity burden was associated with increased odds of nonadherence to adjuvant capecitabine. | Amlani et al., 2016 |
| **Higher number of concomitant medications** | $\geq 4$ concomitant medications vs $\leq 3$ | More concomitant drugs were associated with higher odds of nonadherence in an adjuvant capecitabine cohort. | Amlani et al., 2016 |

**Table 2B.**
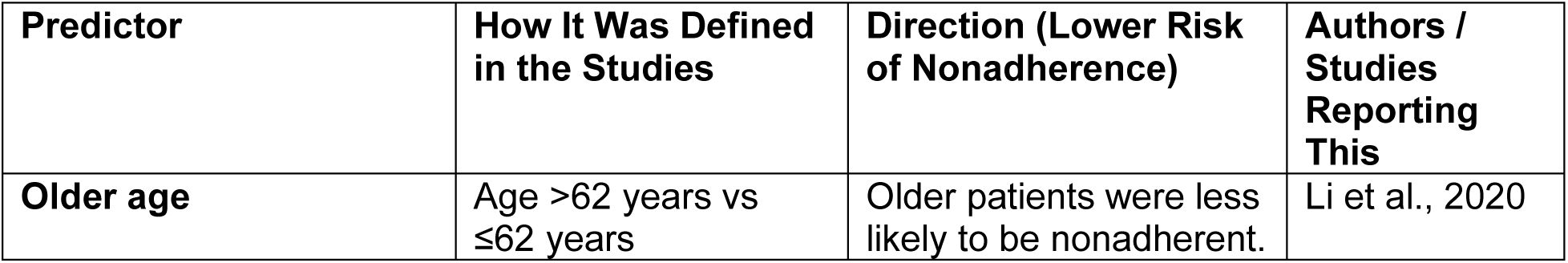
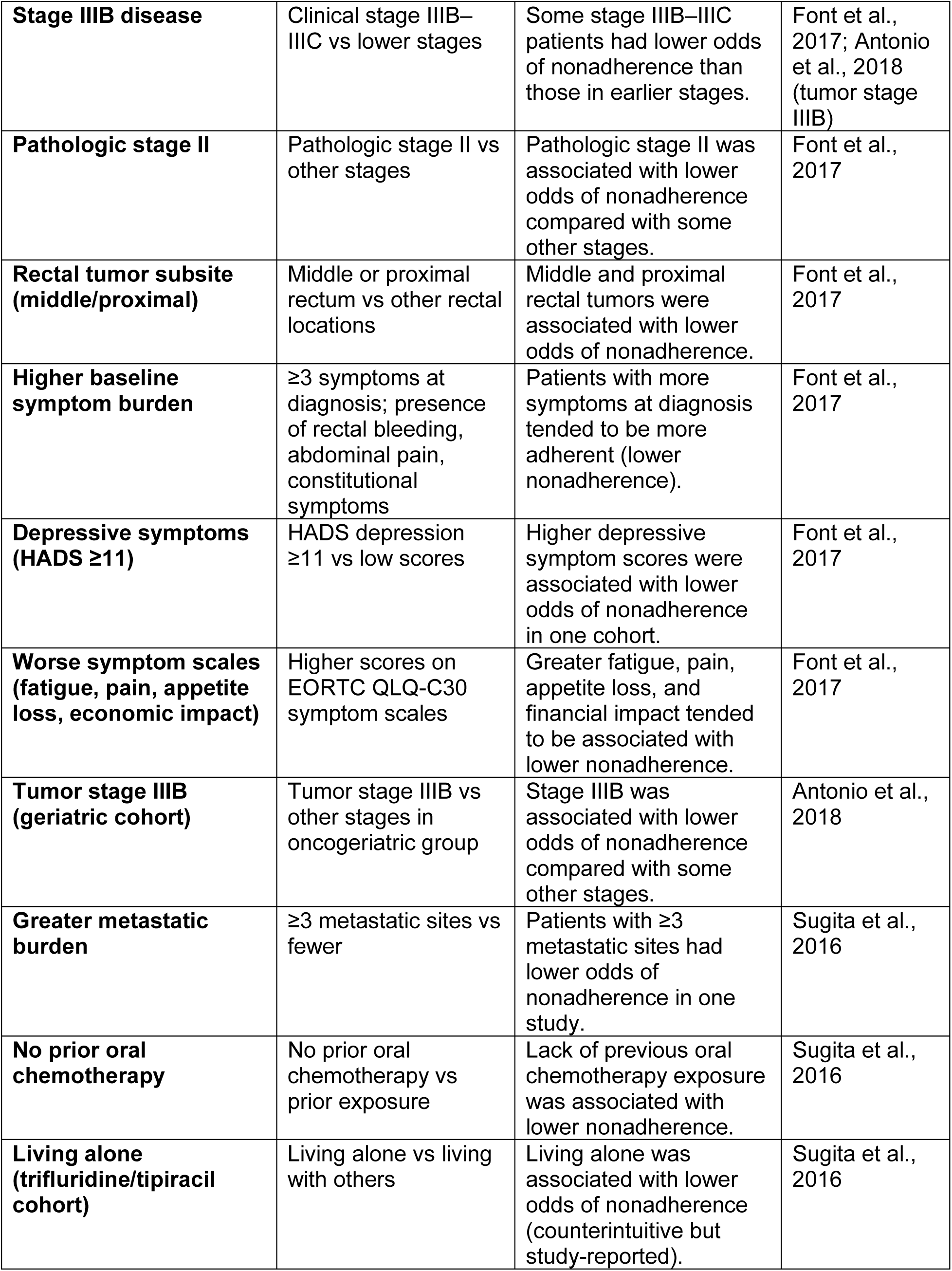
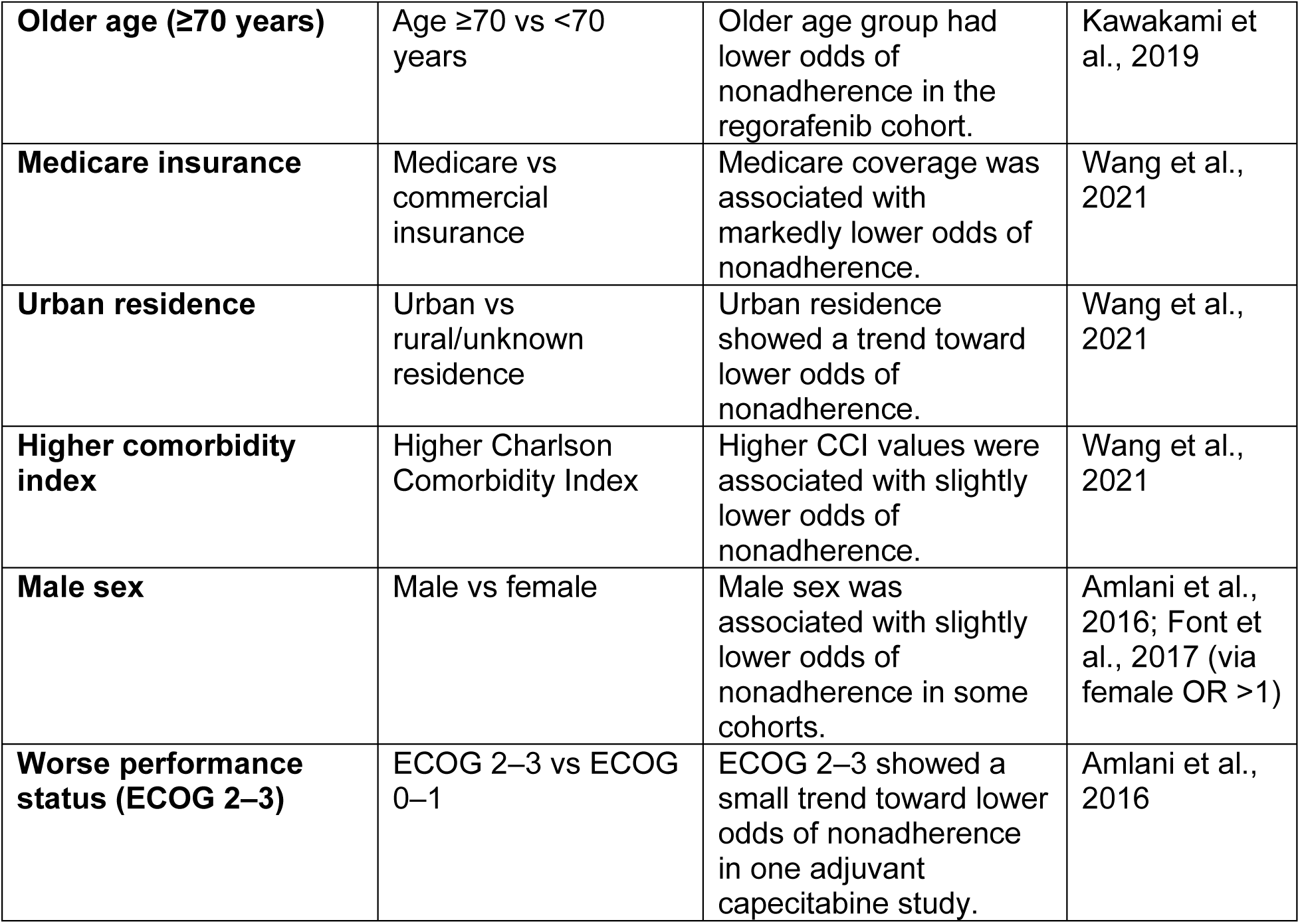
Predictors Associated With Lower Risk of Nonadherence to Colorectal Cancer Chemotherapy (Negative Associations)

| **Predictor** | **How It Was Defined in the Studies** | **Direction (Lower Risk of Nonadherence)** | **Authors / Studies Reporting This** |
| --- | --- | --- | --- |
| **Older age** | Age $>62$ years vs $\leq 62$ years | Older patients were less likely to be nonadherent. | Li et al., 2020 |
| **Stage IIIB disease** | Clinical stage IIIB–IIIC vs lower stages | Some stage IIIB–IIIC patients had lower odds of nonadherence than those in earlier stages. | Font et al., 2017; Antonio et al., 2018 (tumor stage IIIB) |
| **Pathologic stage II** | Pathologic stage II vs other stages | Pathologic stage II was associated with lower odds of nonadherence compared with some other stages. | Font et al., 2017 |
| **Rectal tumor subsite (middle/proximal)** | Middle or proximal rectum vs other rectal locations | Middle and proximal rectal tumors were associated with lower odds of nonadherence. | Font et al., 2017 |
| **Higher baseline symptom burden** | ≥3 symptoms at diagnosis; presence of rectal bleeding, abdominal pain, constitutional symptoms | Patients with more symptoms at diagnosis tended to be more adherent (lower nonadherence). | Font et al., 2017 |
| **Depressive symptoms (HADS ≥11)** | HADS depression ≥11 vs low scores | Higher depressive symptom scores were associated with lower odds of nonadherence in one cohort. | Font et al., 2017 |
| **Worse symptom scales (fatigue, pain, appetite loss, economic impact)** | Higher scores on EORTC QLQ-C30 symptom scales | Greater fatigue, pain, appetite loss, and financial impact tended to be associated with lower nonadherence. | Font et al., 2017 |
| **Tumor stage IIIB (geriatric cohort)** | Tumor stage IIIB vs other stages in oncogeriatric group | Stage IIIB was associated with lower odds of nonadherence compared with some other stages. | Antonio et al., 2018 |
| **Greater metastatic burden** | ≥3 metastatic sites vs fewer | Patients with ≥3 metastatic sites had lower odds of nonadherence in one study. | Sugita et al., 2016 |
| **No prior oral chemotherapy** | No prior oral chemotherapy vs prior exposure | Lack of previous oral chemotherapy exposure was associated with lower nonadherence. | Sugita et al., 2016 |
| **Living alone (trifluridine/tipiracil cohort)** | Living alone vs living with others | Living alone was associated with lower odds of nonadherence (counterintuitive but study-reported). | Sugita et al., 2016 |
| **Older age (≥70 years)** | Age ≥70 vs <70 years | Older age group had lower odds of nonadherence in the regorafenib cohort. | Kawakami et al., 2019 |
| **Medicare insurance** | Medicare vs commercial insurance | Medicare coverage was associated with markedly lower odds of nonadherence. | Wang et al., 2021 |
| **Urban residence** | Urban vs rural/unknown residence | Urban residence showed a trend toward lower odds of nonadherence. | Wang et al., 2021 |
| **Higher comorbidity index** | Higher Charlson Comorbidity Index | Higher CCI values were associated with slightly lower odds of nonadherence. | Wang et al., 2021 |
| **Male sex** | Male vs female | Male sex was associated with slightly lower odds of nonadherence in some cohorts. | Amlani et al., 2016; Font et al., 2017 (via female OR >1) |
| **Worse performance status (ECOG 2–3)** | ECOG 2–3 vs ECOG 0–1 | ECOG 2–3 showed a small trend toward lower odds of nonadherence in one adjuvant capecitabine study. | Amlani et al., 2016 |

**Table 2.**
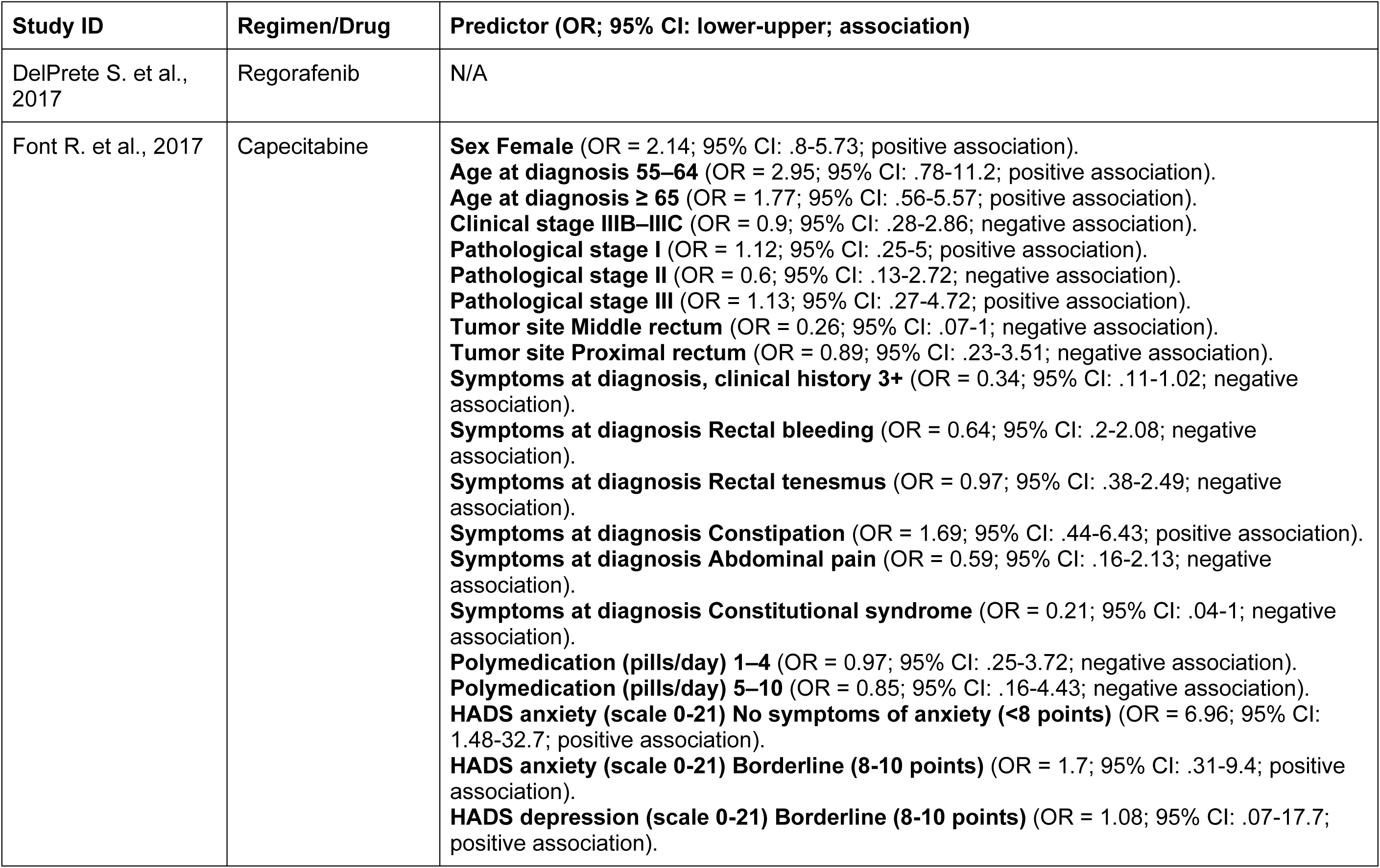
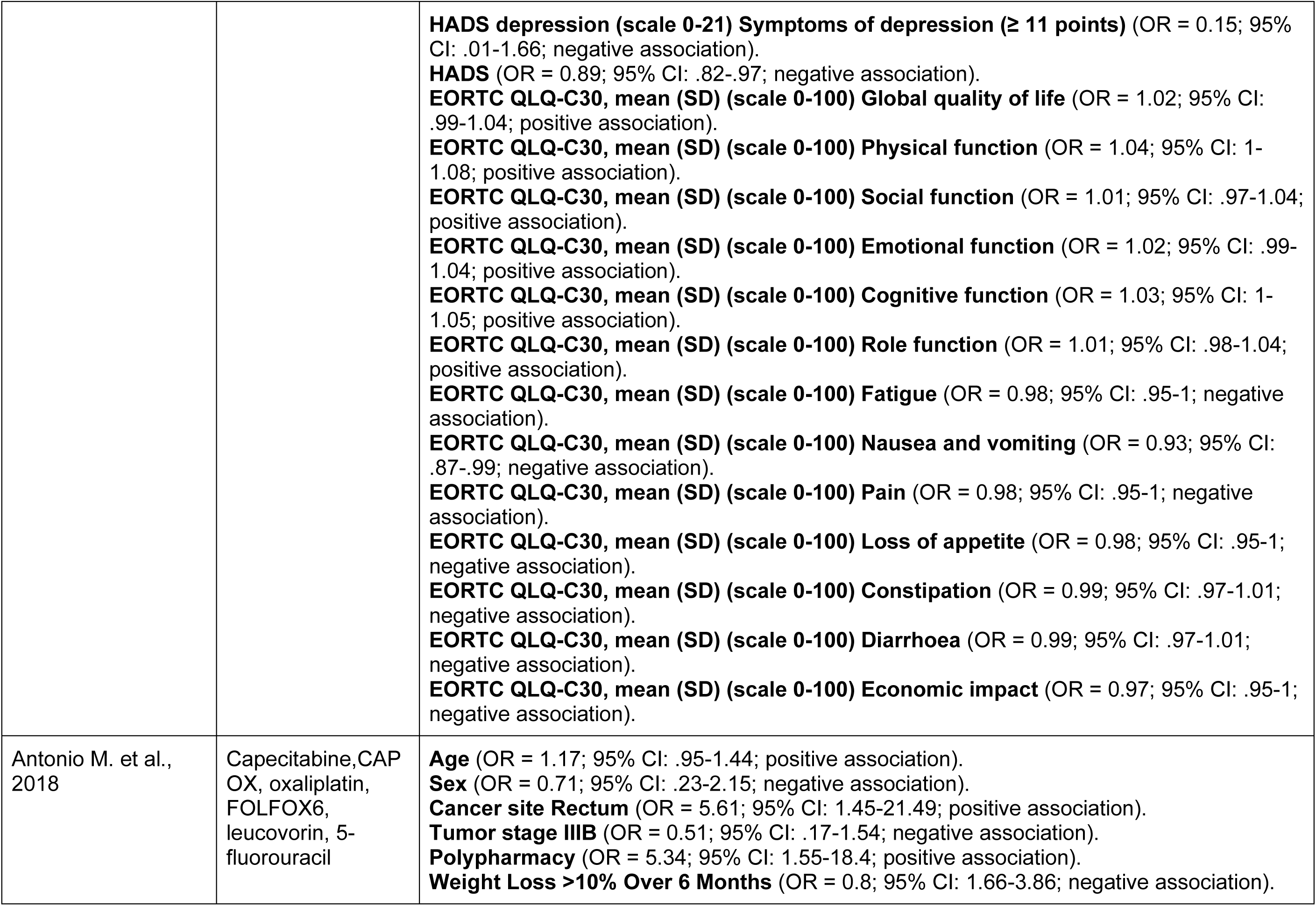
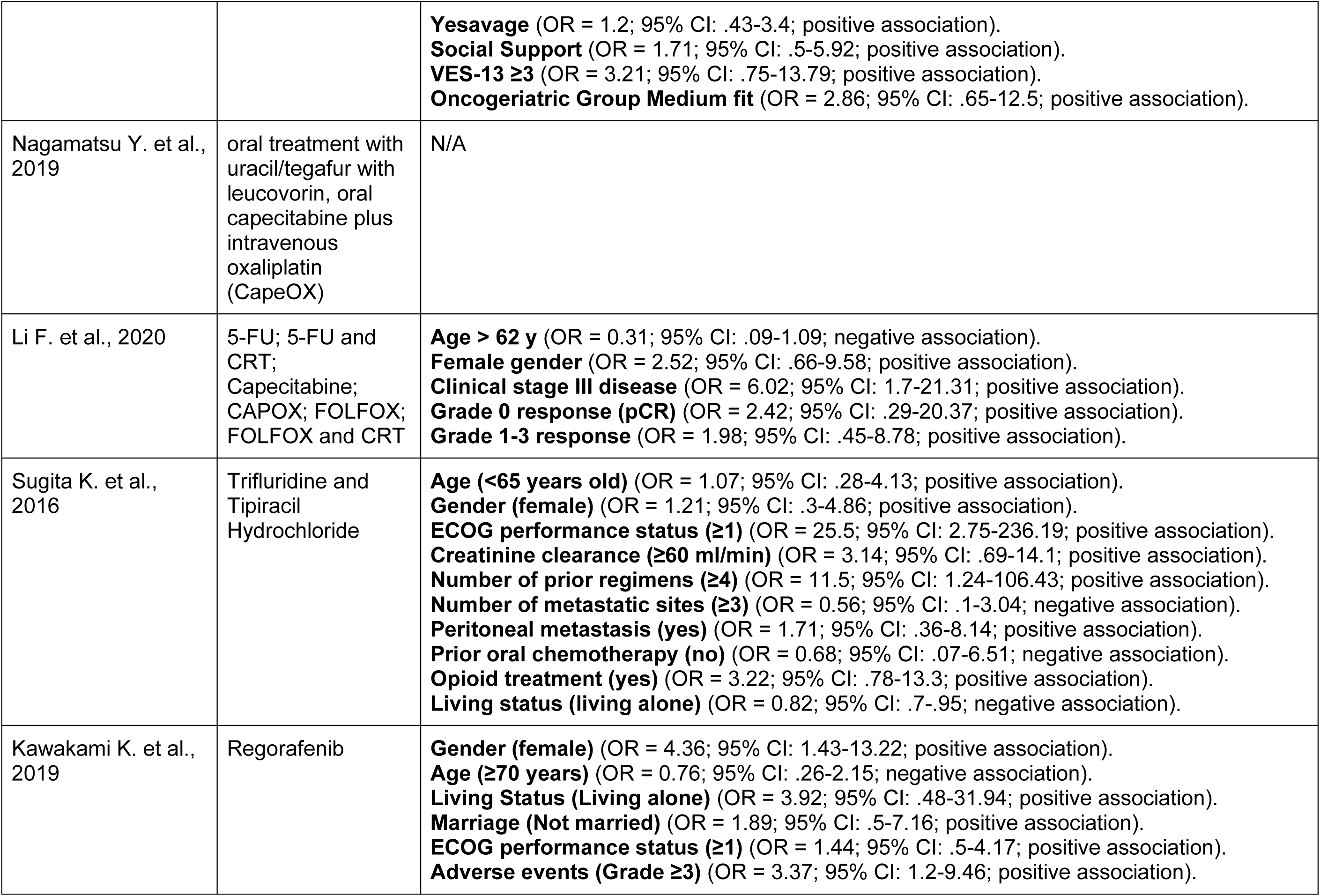
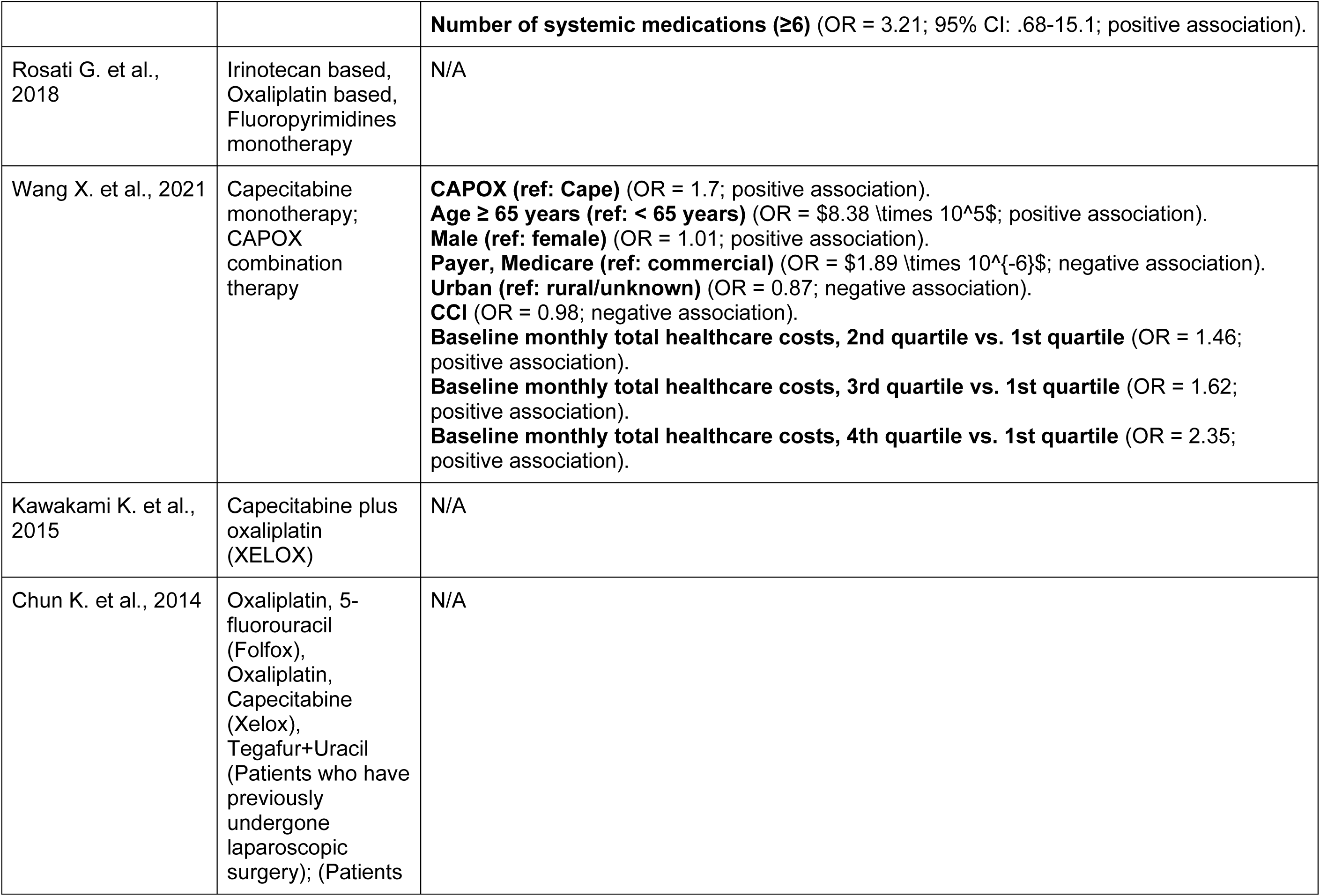
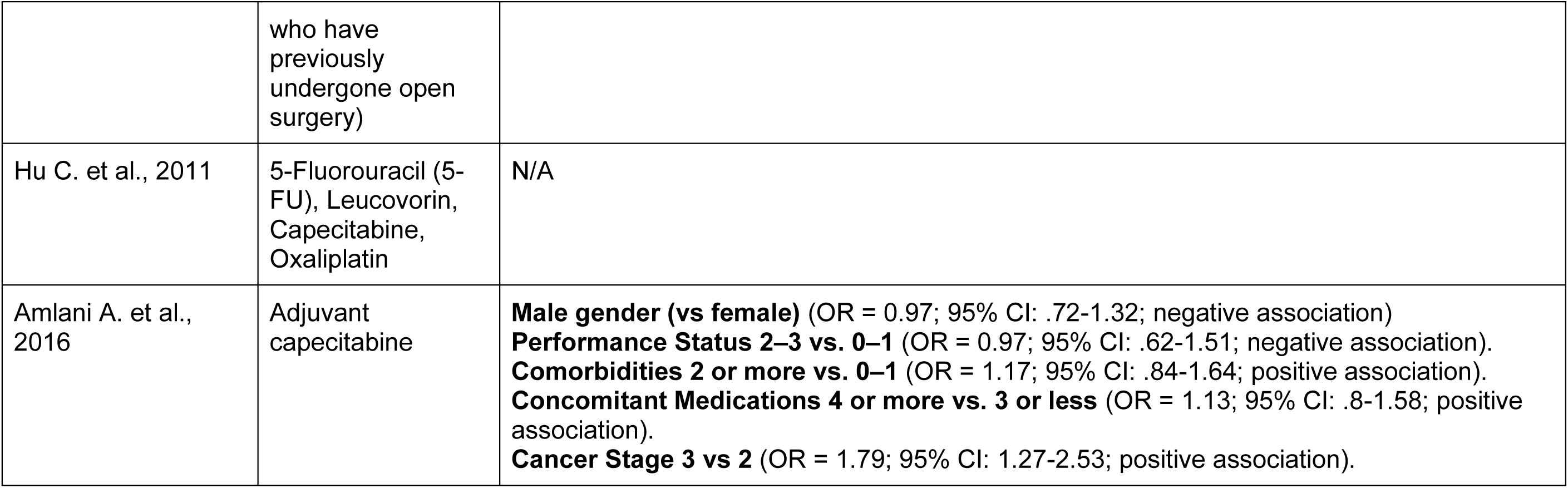
Significant Predictors of Nonadherence.

| Study ID | Regimen/Drug | Predictor (OR; 95% CI: lower-upper; association) |
| --- | --- | --- |
| DelPrete S. et al., 2017 | Regorafenib | N/A |
| Font R. et al., 2017 | Capecitabine | **Sex Female** (OR = 2.14; 95% CI: .8-5.73; positive association).<br>**Age at diagnosis 55–64** (OR = 2.95; 95% CI: .78-11.2; positive association).<br>**Age at diagnosis ≥ 65** (OR = 1.77; 95% CI: .56-5.57; positive association).<br>**Clinical stage IIIB–IIIC** (OR = 0.9; 95% CI: .28-2.86; negative association).<br>**Pathological stage I** (OR = 1.12; 95% CI: .25-5; positive association).<br>**Pathological stage II** (OR = 0.6; 95% CI: .13-2.72; negative association).<br>**Pathological stage III** (OR = 1.13; 95% CI: .27-4.72; positive association).<br>**Tumor site Middle rectum** (OR = 0.26; 95% CI: .07-1; negative association).<br>**Tumor site Proximal rectum** (OR = 0.89; 95% CI: .23-3.51; negative association).<br>**Symptoms at diagnosis, clinical history 3+** (OR = 0.34; 95% CI: .11-1.02; negative association).<br>**Symptoms at diagnosis Rectal bleeding** (OR = 0.64; 95% CI: .2-2.08; negative association).<br>**Symptoms at diagnosis Rectal tenesmus** (OR = 0.97; 95% CI: .38-2.49; negative association).<br>**Symptoms at diagnosis Constipation** (OR = 1.69; 95% CI: .44-6.43; positive association).<br>**Symptoms at diagnosis Abdominal pain** (OR = 0.59; 95% CI: .16-2.13; negative association).<br>**Symptoms at diagnosis Constitutional syndrome** (OR = 0.21; 95% CI: .04-1; negative association).<br>**Polymedication (pills/day) 1–4** (OR = 0.97; 95% CI: .25-3.72; negative association).<br>**Polymedication (pills/day) 5–10** (OR = 0.85; 95% CI: .16-4.43; negative association).<br>**HADS anxiety (scale 0-21) No symptoms of anxiety (<8 points)** (OR = 6.96; 95% CI: 1.48-32.7; positive association).<br>**HADS anxiety (scale 0-21) Borderline (8-10 points)** (OR = 1.7; 95% CI: .31-9.4; positive association).<br>**HADS depression (scale 0-21) Borderline (8-10 points)** (OR = 1.08; 95% CI: .07-17.7; positive association). |
|  |  | **HADS depression (scale 0-21) Symptoms of depression ($\geq 11$ points)** (OR = 0.15; 95% CI: .01-1.66; negative association).<br>**HADS** (OR = 0.89; 95% CI: .82-.97; negative association).<br>**EORTC QLQ-C30, mean (SD) (scale 0-100) Global quality of life** (OR = 1.02; 95% CI: .99-1.04; positive association).<br>**EORTC QLQ-C30, mean (SD) (scale 0-100) Physical function** (OR = 1.04; 95% CI: 1-1.08; positive association).<br>**EORTC QLQ-C30, mean (SD) (scale 0-100) Social function** (OR = 1.01; 95% CI: .97-1.04; positive association).<br>**EORTC QLQ-C30, mean (SD) (scale 0-100) Emotional function** (OR = 1.02; 95% CI: .99-1.04; positive association).<br>**EORTC QLQ-C30, mean (SD) (scale 0-100) Cognitive function** (OR = 1.03; 95% CI: 1-1.05; positive association).<br>**EORTC QLQ-C30, mean (SD) (scale 0-100) Role function** (OR = 1.01; 95% CI: .98-1.04; positive association).<br>**EORTC QLQ-C30, mean (SD) (scale 0-100) Fatigue** (OR = 0.98; 95% CI: .95-1; negative association).<br>**EORTC QLQ-C30, mean (SD) (scale 0-100) Nausea and vomiting** (OR = 0.93; 95% CI: .87-.99; negative association).<br>**EORTC QLQ-C30, mean (SD) (scale 0-100) Pain** (OR = 0.98; 95% CI: .95-1; negative association).<br>**EORTC QLQ-C30, mean (SD) (scale 0-100) Loss of appetite** (OR = 0.98; 95% CI: .95-1; negative association).<br>**EORTC QLQ-C30, mean (SD) (scale 0-100) Constipation** (OR = 0.99; 95% CI: .97-1.01; negative association).<br>**EORTC QLQ-C30, mean (SD) (scale 0-100) Diarrhoea** (OR = 0.99; 95% CI: .97-1.01; negative association).<br>**EORTC QLQ-C30, mean (SD) (scale 0-100) Economic impact** (OR = 0.97; 95% CI: .95-1; negative association). |
| Antonio M. et al., 2018 | Capecitabine,CAP<br>OX, oxaliplatin,<br>FOLFOX6,<br>leucovorin, 5-fluorouracil | **Age** (OR = 1.17; 95% CI: .95-1.44; positive association).<br>**Sex** (OR = 0.71; 95% CI: .23-2.15; negative association).<br>**Cancer site Rectum** (OR = 5.61; 95% CI: 1.45-21.49; positive association).<br>**Tumor stage IIIB** (OR = 0.51; 95% CI: .17-1.54; negative association).<br>**Polypharmacy** (OR = 5.34; 95% CI: 1.55-18.4; positive association).<br>**Weight Loss >10% Over 6 Months** (OR = 0.8; 95% CI: 1.66-3.86; negative association). |
|  |  | **Yesavage** (OR = 1.2; 95% CI: .43-3.4; positive association).<br>**Social Support** (OR = 1.71; 95% CI: .5-5.92; positive association).<br>**VES-13 $\geq 3$** (OR = 3.21; 95% CI: .75-13.79; positive association).<br>**Oncogeriatric Group Medium fit** (OR = 2.86; 95% CI: .65-12.5; positive association). |
| Nagamatsu Y. et al., 2019 | oral treatment with uracil/tegafur with leucovorin, oral capecitabine plus intravenous oxaliplatin (CapeOX) | N/A |
| Li F. et al., 2020 | 5-FU; 5-FU and CRT;<br>Capecitabine;<br>CAPOX; FOLFOX;<br>FOLFOX and CRT | **Age > 62 y** (OR = 0.31; 95% CI: .09-1.09; negative association).<br>**Female gender** (OR = 2.52; 95% CI: .66-9.58; positive association).<br>**Clinical stage III disease** (OR = 6.02; 95% CI: 1.7-21.31; positive association).<br>**Grade 0 response (pCR)** (OR = 2.42; 95% CI: .29-20.37; positive association).<br>**Grade 1-3 response** (OR = 1.98; 95% CI: .45-8.78; positive association). |
| Sugita K. et al., 2016 | Trifluridine and Tipiracil Hydrochloride | **Age (<65 years old)** (OR = 1.07; 95% CI: .28-4.13; positive association).<br>**Gender (female)** (OR = 1.21; 95% CI: .3-4.86; positive association).<br>**ECOG performance status ($\geq 1$)** (OR = 25.5; 95% CI: 2.75-236.19; positive association).<br>**Creatinine clearance ($\geq 60$ ml/min)** (OR = 3.14; 95% CI: .69-14.1; positive association).<br>**Number of prior regimens ($\geq 4$)** (OR = 11.5; 95% CI: 1.24-106.43; positive association).<br>**Number of metastatic sites ($\geq 3$)** (OR = 0.56; 95% CI: .1-3.04; negative association).<br>**Peritoneal metastasis (yes)** (OR = 1.71; 95% CI: .36-8.14; positive association).<br>**Prior oral chemotherapy (no)** (OR = 0.68; 95% CI: .07-6.51; negative association).<br>**Opioid treatment (yes)** (OR = 3.22; 95% CI: .78-13.3; positive association).<br>**Living status (living alone)** (OR = 0.82; 95% CI: .7-.95; negative association). |
| Kawakami K. et al., 2019 | Regorafenib | **Gender (female)** (OR = 4.36; 95% CI: 1.43-13.22; positive association).<br>**Age ($\geq 70$ years)** (OR = 0.76; 95% CI: .26-2.15; negative association).<br>**Living Status (Living alone)** (OR = 3.92; 95% CI: .48-31.94; positive association).<br>**Marriage (Not married)** (OR = 1.89; 95% CI: .5-7.16; positive association).<br>**ECOG performance status ($\geq 1$)** (OR = 1.44; 95% CI: .5-4.17; positive association).<br>**Adverse events (Grade $\geq 3$)** (OR = 3.37; 95% CI: 1.2-9.46; positive association). |
|  |  | **Number of systemic medications (≥6)** (OR = 3.21; 95% CI: .68-15.1; positive association). |
| Rosati G. et al., 2018 | Irinotecan based,<br>Oxaliplatin based,<br>Fluoropyrimidines<br>monotherapy | N/A |
| Wang X. et al., 2021 | Capecitabine<br>monotherapy;<br>CAPOX<br>combination<br>therapy | **CAPOX (ref: Cape)** (OR = 1.7; positive association).<br>**Age ≥ 65 years (ref: < 65 years)** (OR = \$8.38 \times 10^5\$; positive association).<br>**Male (ref: female)** (OR = 1.01; positive association).<br>**Payer, Medicare (ref: commercial)** (OR = \$1.89 \times 10^{-6}\$; negative association).<br>**Urban (ref: rural/unknown)** (OR = 0.87; negative association).<br>**CCI** (OR = 0.98; negative association).<br>**Baseline monthly total healthcare costs, 2nd quartile vs. 1st quartile** (OR = 1.46; positive association).<br>**Baseline monthly total healthcare costs, 3rd quartile vs. 1st quartile** (OR = 1.62; positive association).<br>**Baseline monthly total healthcare costs, 4th quartile vs. 1st quartile** (OR = 2.35; positive association). |
| Kawakami K. et al., 2015 | Capecitabine plus<br>oxaliplatin<br>(XELOX) | N/A |
| Chun K. et al., 2014 | Oxaliplatin, 5-fluorouracil<br>(Folfox),<br>Oxaliplatin,<br>Capecitabine<br>(Xelox),<br>Tegafur+Uracil<br>(Patients who have<br>previously<br>undergone<br>laparoscopic<br>surgery); (Patients | N/A |
|  | who have previously undergone open surgery) |  |
| Hu C. et al., 2011 | 5-Fluorouracil (5-FU), Leucovorin, Capecitabine, Oxaliplatin | N/A |
| Amlani A. et al., 2016 | Adjuvant capecitabine | **Male gender (vs female)** (OR = 0.97; 95% CI: .72-1.32; negative association)<br>**Performance Status 2–3 vs. 0–1** (OR = 0.97; 95% CI: .62-1.51; negative association).<br>**Comorbidities 2 or more vs. 0–1** (OR = 1.17; 95% CI: .84-1.64; positive association).<br>**Concomitant Medications 4 or more vs. 3 or less** (OR = 1.13; 95% CI: .8-1.58; positive association).<br>**Cancer Stage 3 vs 2** (OR = 1.79; 95% CI: 1.27-2.53; positive association). |

**Table 3.** Risk of Bias Assessment.

| **Study ID** | **JBI Score** | **Study quality** |
| --- | --- | --- |
| DelPrete S. et al., 2017 | 77.78% | Moderate |
| Font R. et al., 2017 | 88.89% | High |
| Antonio M. et al., 2018 | 77.78% | Moderate |
| Nagamatsu Y. et al., 2019 | 66.67% | Moderate |
| Li F. et al., 2020 | 66.67% | Moderate |
| Sugita K. et al., 2016 | 66.67% | Moderate |
| Kawakami K. et al., 2019 | 100% | High |
| Rosati G. et al., 2018 | 62.50% | Moderate |
| Wang X. et al., 2021 | 77.78% | Moderate |
| Kawakami K. et al., 2015 | 33.33% | Low |
| Chun K. et al., 2014 | 50.00% | Low |
| Hu C. et al., 2011 | 77.78% | Moderate |
| Amlani A. et al., 2016 | 88.89% | High |

## DISCUSSION

### Principal Findings

In studies primarily utilizing administrative or claims data to calculate adherence, the pooled adherence rate was 40%. In studies primarily utilizing data from patient medical charts, physician notes, or pharmacist verification of treatment cycles/doses completed to calculate adherence or in studies primarily utilizing data from patient questionnaires, direct pill counts, or patient self-report to calculate adherence, the pooled adherence rate was 83%. Predictors associated with a higher risk (positive association) include Cancer Stage 3 vs 2, ECOG performance status (≥1), Number of prior regimens (≥4), HADS anxiety (No symptoms of anxiety <8 points), Clinical stage III disease, Female gender, Adverse events (Grade ≥3), Cancer site Rectum, and Polypharmacy. Predictors associated with a lower risk (negative association) are Living status (living alone), HADS (continuous measure), and Nausea and vomiting (EORTC QLQ-C30).

### Potential Causes for Statistically Significant Predictors of Higher Odds of Non-Adherence

Several factors contribute to statistically significant predictors of higher odds of non-adherence. For “Cancer Stage (Stage 3 vs. 2; Clinical Stage III),^9, 17^ advanced diseases can lead to increased symptom burden, psychological distress,^18^ or a perception of treatment ineffectiveness,^19^ coupled with more complex regimens, increasing opportunities for errors. “ECOG Performance Status of ≥1”^10^ indicates greater physical limitations, fatigue, and reduced functional capacity, often with co-occurring cognitive impairment, complicating self-management. ^20^ Patients with “Four or More Prior Regimens”^10^ may experience “treatment fatigue” and cumulative side effects, leading to burnout.^21^ “Primary School Education Only”^5^ often correlates with lower health literacy, impeding comprehension of complex medical instructions.^22^ The “CAPOX Regimen vs. Capecitabine”^13^ is associated with higher incidence and severity of side effects, potentially leading to intentional dose modifications.^23^ “Higher Baseline Monthly Total Healthcare Costs”^13^ can force patients to ration or delay medications due to financial burden. “Female Gender”^11^ is a complex predictor, possibly involving higher rates of depression/anxiety,^24^ specific symptom experiences, or differing caregiving burdens impacting self-care. ^25^ Lastly, “Experiencing Grade ≥3 Adverse Events”^11^ directly impacts quality of life, often leading to intentional non-adherence as patients seek to alleviate discomfort.^26^

### Implications of Results for Practice, Policy, and Future Research

A range of interventional methods can address these predictors of non-adherence. For “Cancer Stage,” patient navigation programs offer tailored support, education, and care coordination, improving adherence in complex pathways.^27^ Comprehensive nursing and pharmacy support addresses non-adherence related to “ECOG Performance Status of ≥1” through detailed education, proactive symptom management, and regular follow-up.^28^ Psychosocial interventions and fatigue management, including CBT and strategies for cancer-related fatigue, are beneficial for patients with “Four or More Prior Regimens.”^29^ For those with “Primary School Education Only,” strategies like using plain language, visual aids, and culturally appropriate materials enhance health literacy and comprehension.^30^ Proactive side effect management, involving timely care team intervention and digital health tools, supports patients experiencing severe side effects from regimens like “CAPOX vs. Capecitabine.”^31^ To mitigate the impact of “Higher Baseline Monthly Total Healthcare Costs”, interventions include screening for financial distress, connecting patients with financial counselors, and assisting with patient assistance programs.^32^ For “Female Gender,” tailored psychosocial support and screening can manage psychological distress (e.g., depression, anxiety), and support groups or counseling specific to women’s experiences with cancer can be helpful.^33, 34^ Finally, proactive symptom management through comprehensive pre-treatment education, timely assessment of adverse events, and nurse-led follow-up calls addresses non-adherence when “Experiencing Grade ≥3 Adverse Events.” ^35^ Future research can improve our understanding of the effectiveness of these interventions on improving patient adherence to oral colorectal cancer medications, and guide healthcare policy development in this area.

### Strengths and Limitations

Strengths: Conducting separate random-effects meta-analyses for each adherence measurement subgroup allowed for robust pooled estimates and assessment of statistical heterogeneity using the I^2^ statistic. The use of the Joanna Briggs Institute Critical Appraisal Checklist for Cohort Studies provided a standardized and validated framework for assessing study quality and risk of bias.

Limitations: High heterogeneity across the original studies, with widely varying methodologies and adherence definitions, made direct comparisons of reported adherence rates complex. This review included studies from only four databases. Non-English studies were excluded.

## Data Availability

All data produced in the present work are contained in the manuscript

## Acknowledgements of research funding support for the study

New York University Martin Luther King Jr. Program Summer Undergraduate Research Funding Ohio State University College of Pharmacy Summer Undergraduate Research Funding

## Other Information

### Registration and Protocol

The review was not registered. A protocol was not prepared.

### Availability of data, code, and other materials

Data extracted from included studies can be identified in the Results section.

